# Epidemiology of malaria in an area with pyrethroid-resistant vectors in south-western Burkina Faso: a pre-intervention study

**DOI:** 10.1101/2020.06.04.20120105

**Authors:** Anthony Somé, Issaka Zongo, Bertin N’cho Tchiekoi, Dieudonné D. Soma, Barnabas Zogo, Mamadou Ouattara, Anyirékun F. Somé, Amal Dahounto, Alphonsine A. Koffi, Cédric Pennetier, Nicolas Moiroux, Seni Kouanda, Roch K. Dabiré

**Author notes:** **Corresponding author:** Anthony Somé.

## Abstract

**Background:** The objective of this study was to update malaria epidemiological profile prior to the implementation of a randomized controlled trial aiming to evaluate the efficacy of new vector control tools in complementary to the use of long-lasting insecticidal nets in Burkina Faso.

**Methods:** We carried out active and passive cross-sectional surveys to estimate the prevalence and incidence of malaria infection from August 2016 to July 2017 in 27 villages of the Diebougou health district.

**Results:** With the passive survey, we extracted data from 4814 patients included in the study from August 2016 to July 2017. Malaria incidence showed a seasonal distribution, with an overall incidence rate estimated at 414.3 per 1000 person-years. In the active cross sectional surveys, we enrolled 2839, 2594 and 2337 participants respectively in September 2016, December 2016 and June 2017. Prevalence of malaria infection were respectively 41.5%, 43.5% and 32.3% in September 2016, in December 2016 and June 2017. Multivariate analysis showed that girls seemed to have a lower risk of malaria infection (OR = 0.86; 95% CI = 0.79 - 0.95; *p* = 0.004). The risk of malaria infection was significantly lower in third survey (June 2017) at the beginning of the rainy season (OR = 0.69; 95% CI = 0.6 - 0.8; *p* < 0.001) compared to the first survey (September 2016) which was performed during the rainy season. Children aged 6 to 59 months had a higher risk of malaria infection compared to those aged 10 to 17 years (OR = 0.58; 95% CI = 0.51 - 0.66).

**Conclusion:** Malaria burden remains high in this region of Burkina Faso despite substantial efforts made in malaria control during this current decade. Children under 5 years old were subject of malaria burden in this setting. This results reinforce the urgent need to develop alternative control strategies to complement those already existing.

## Background

Malaria remains a major cause of morbidity and mortality in sub-Saharan Africa, especially in children under the age of 5 [1]. In 2017, approximately 198 million malaria cases occurred globally worldwide, causing 584000 deaths. Most cases (90%) occurred in Africa, and most deaths (78%) were in children under 5 years old [2].

In Burkina Faso, malaria is the main cause of consultation, hospitalization and mortality in health centers especially within children under 5 years’ age and pregnant women. Indeed in 2017, 43.5% of consultations, 60.5% of hospitalizations and 35.9% of deaths in primary health centers (PHCs) in the country were due to malaria [3].

Many countries in the recent years have made significant progress in preventing malaria by largely focusing on vector control through LLINs and indoor residual spraying (IRS) of insecticides [4]. It is estimated that between 2000 and 2010, LLINs has saved more than 908000 lives, and since 2006, prevented three-quarters of deaths due to malaria [5].

However, the widespread use of LLINs leads to the development of vector resistance to insecticide. This insecticide resistance can reduce the effectiveness of these interventions and perhaps reverse progress in reducing malaria morbidity [6]. New strategies must be developed to reduce the development and spread of insecticide resistance and preserve the effectiveness of currently available insecticides and malaria control interventions. It is obvious that increasing the level of resistance corresponds to a decrease in the effectiveness of vector control strategies implementation [7].

As part of resistance management, the World Health Organization (WHO) recommends the development of alternative control strategies to complement those already existing and recognized for their effectiveness, but threatened by the expansion of resistance, in order to limit this expansion and maintain effectiveness [4]. The present study is part of a randomized controlled trial (RCT) in the framework of a project called REACT designed to investigate whether the combination of LLINs with other vector control strategies in areas with pyrethroid resistance can provide additional protection against malaria. Updated malaria epidemiology data in endemic areas are needed to serve as baseline to evaluate public health impact of alternative interventions implemented and to monitor effects of malaria control activities over time. We sought to describe the epidemiology of malaria prior to the conduct of the RCT. Specifically, in this paper, we assessed the incidence and prevalence of malaria during the rainy and dry seasons in Diebougou health district in the context of widespread insecticide resistance.

## Materials and methods

### Study area

This study was part of a randomized controlled trial carried out in 27 villages in Diebougou health district from August 2016 to July 2017. These villages were selected based on geographical (distance between two villages higher than 2 km and accessibility during the rainy season) and demographic (a population size ranging from 200 to 500 inhabitants) criteria. The Diebougou health district is located in South-Western Burkina Faso, in the province of Bougouriba (**Figure 1**). With a total area of 2774 Km^2^, it covers five departments namely Diebougou, Tiankoura, Dolo, Iolonioro, and Bondigui and 154 villages. The district has fourteen PHCs and one district hospital. In 2016, the district served a total 135740 inhabitants comprised of 51.99% females. Most of its population is rural (85.42%) with 32% of children under five years [3]. The climate is a sub-Sudan type with a rainy season of six months from May to October and a dry season from November to April. The population is composed of several ethnic groups, the main ones being Djan, Dagara, Birifor, Lobi, Pougouli, Dogossè, Mossi and Fulani [8].

**Figure 1.**
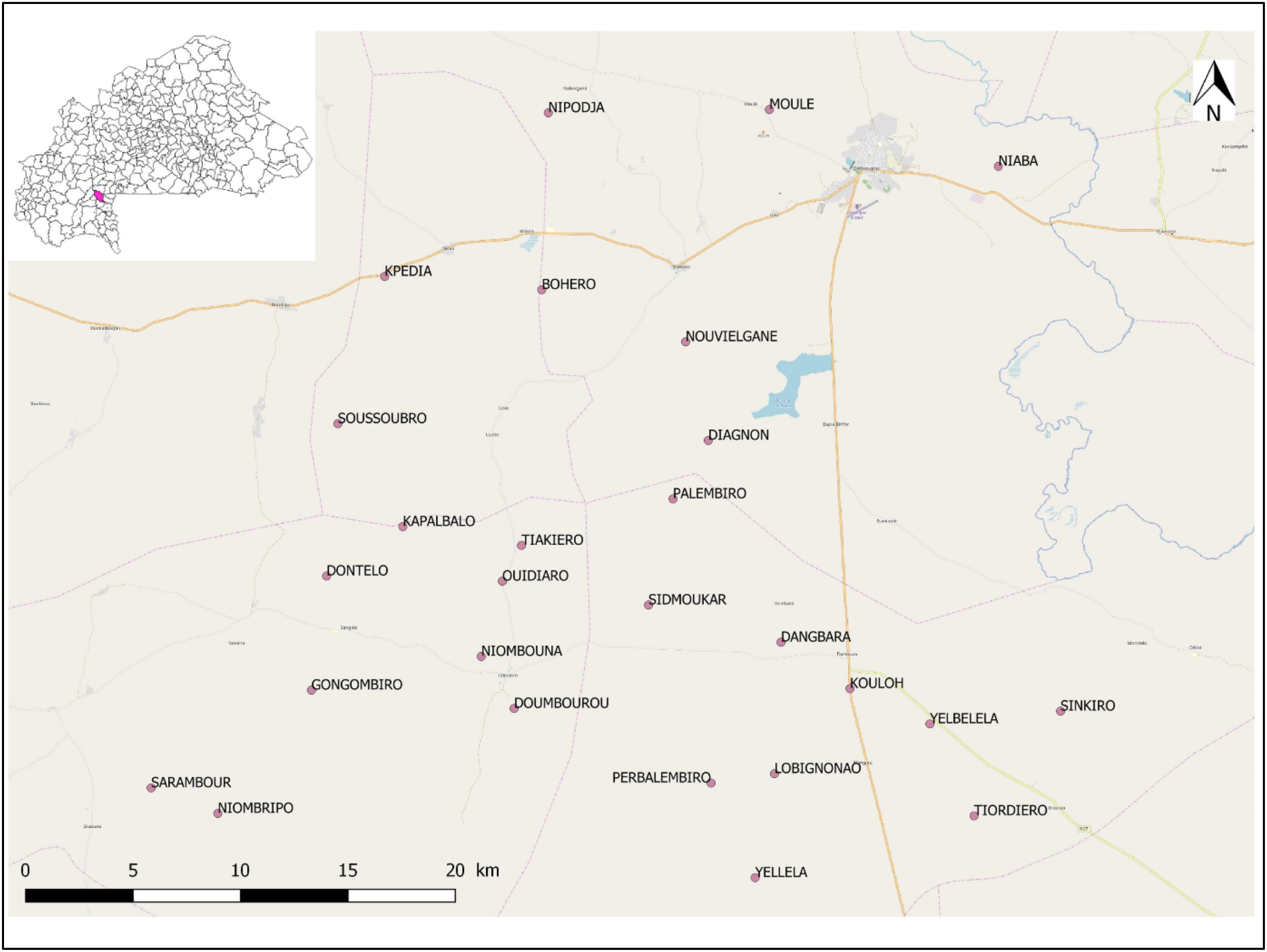
Study area with the 27 target villages.

### Study population

A census was carried out in august 2016 in each of these 27 villages. Houses were numbered, georeferenced, and information on the household’s residents (name, sex, age, level of education, profession and relationship with the household head) were collected. All inhabitants of the village were enrolled for passive cases detection while inclusion criteria in the active surveys were: age between 6 months and 17 years, residence in one of the target villages, provision of written informed consent or assent.

### Study design and procedure

We combined fixed cross-sectional surveys directly in the selected villages and passive cases detection in primary health centers covering the selected villages. All surveys were completed over one year (August 2016 to July 2017).

#### Passive malaria cases detection

The passive detection was carried out in thirteen PHCs covering the twenty-seven villages and each village depended on a single primary health center. A dedicated study nurse visited each primary health center once every two months to review all outpatient and inpatient data in registers retrospectively. All the individuals residents in the selected villages and visiting these health centers were enrolled in the study. All consultants irrespective of conditions, age or sex were recorded. Variables recorded included the village where the subject came from, the age, sex, reason of visit (symptoms), temperature, diagnostic, rapid diagnostic test (RDT) for malaria done or not, RDT results (positive or negative or undetermined), treatment prescribed. The diagnosis of malaria was based on the association of fever (or fever-related symptoms) with a positive RDT in all primary health centers. These data are then checked and uploaded on a server for further analysis.

#### Active malaria cases detection

Three cross-sectional surveys were carried out in all of the 27 target villages to estimate malaria prevalence: the first survey in September 2016 during the rainy season, the second in December 2016 during the dry season and the last in June 2017 at the beginning of the rainy season. On a fixed period per survey, data were collected by two teams of qualified nurses and laboratory technicians under the supervision of a physician. The population was informed several days in advance of active survey through community health workers and participants were invited to gather in a village square with their legal guardians. The participants were interviewed by the field workers on the socio demographic data and recent history of fever. After this interview, participants benefited a clinical evaluation (physical examination and nutritional parameters measurement) and finger prick for thick and thin smear to estimate parasite densities. In average, two villages were visited per day. During the following surveys, participants who missed the initial census or previous surveys were counted and enrolled in the study.

### Laboratory procedures

Blood smears obtained from all participants by finger prick were air-dried, stained with Giemsa 10% for 10 min and examined by light microscopy by experienced microscopists in the laboratory of IRSS laboratory, Bobo-Dioulasso. Parasite density was determined by counting the number of asexual parasites per 500 white blood cells, assuming a total white blood cells count of 8000/μL. To assure quality of the microscopic examinations, all slides were re-examined by a second microscopist.

### Data analysis

All data were recorded on android tablets using Open Data Kit (ODK) and exported for analysis with STATA version 13 (Stata Corp LP, College Station, TX, USA).

Descriptive statistics were used to summarize baseline values and demographic data. 95% confidence intervals were also calculated. Malaria prevalence was computed by dividing the number of people who showed infection by the total number of people examined. Incidence rates of clinical malaria, for passive cases detection, were calculated as the number of news cases per 1000 populations per year. Possible predictors of clinical malaria at cross-sectional surveys (CSS) were studied using bivariate logistic regression models. Factors with *p*-value < 0.20 were selected for inclusion in a multivariate model constructed using a backwards elimination procedure that retained only factors with a *P* value < 0.05. Statistical significance was assessed using Wald tests at the 5% two-sided significance level.

### Ethics approval and consent to participate

Field workers were trained on the study procedure, informed consent administration procedure and data collection. They were helped in the field by community health workers. The study was approved by the Institutional Ethics Committee for Health Research of the Institut de Recherche en Sciences de la Santé (Reference number N°A06/2016/CEIRES). Written informed consents were obtained from all parents or guardians of children who participated in the study. Participants were explaining the full scoop of the study and their right to participate or not without any prejudice; participation could be stopped at any time without any explanation. Subjects diagnosed with malaria were treated in accordance with the National Malaria Control Program guideline.

## Results

### Socio-demographic characteristics of the study population

At the beginning of the study, the census identified 7337 inhabitants in twenty-seven villages distributed in 1354 households; Females represented 54.2% (3978/7337) of the sample, children under 5 years old accounted for 14.5% (1061/7337) and children aged 6 months to 17 years 54.9% (4028/7337). The most populated village was Sarambour with 489 inhabitants while the least populated was Yelbelela with 160 inhabitants. Socio-demographic characteristics of participants in each cross-sectional survey are summarized in **Table 1**. The passive cases detection survey of malaria reported a total of 4814 patients who sought care in the primary health centers between August 2016 and July 2017. Children aged 6 to 59 months accounted for 55% in the total sample of care seekers. Demographical characteristics of participants to the passive survey are detailed in **Table 2**.

**Table 1.** Socio-demographical characteristics of the study participants of the CSS.

|  | **Sept. 2016**<br>**N (%)** | **Dec. 2016**<br>**N (%)** | **June 2017**<br>**N (%)** | **Total**<br>**N (%)** |
| --- | --- | --- | --- | --- |
| **Number of individuals** | 2839 | 2594 | 2337 | 7770 |
| **Sex** |  |  |  |  |
| Male | 1418 (49.9) | 1284 (49.5) | 1063 (45.5) | 3765 (48.5) |
| Female | 1421 (50.1) | 1310 (50.5) | 1274 (54.5) | 4005 (51.5) |
| **Age group** |  |  |  |  |
| 6 - 59 months | 848 (29.9) | 803 (30.9) | 719 (30.7) | 2370 (30.5) |
| 5 - 9 years | 1080 (38.0) | 1018 (39.3) | 985 (42.2) | 3083 (39.7) |
| 10 - 17 years | 911 (32.1) | 773 (29.8) | 633 (27.1) | 2317 (29.8) |
| **Village** |  |  |  |  |
| Bohero | 116 (4.1) | 128 (4.9) | 114 (4.9) | 358 (4.6) |
| Dangbara | 90 (3.1) | 80 (3.1) | 66 (2.8) | 236 (3.0) |
| Diagno | 91 (3.2) | 79 (3.0) | 63 (2.7) | 233 (2.9) |
| Dombourou | 128 (4.5) | 99 (3.8) | 118 (5.0) | 345 (4.4) |
| Dontélo | 92 (3.2) | 86 (3.3) | 111 (4.7) | 289 (3.7) |
| Gongombiro | 181 (6.4) | 145 (5.6) | 116 (4.9) | 442 (5.7) |
| Kouloh | 101 (3.5) | 104 (4.0) | 96 (4.1) | 301 (3.9) |
| Kpalbalo | 142 (5.0) | 103 (3.9) | 102 (4.3) | 347 (4.5) |
| Kpédia | 131 (4.6) | 129 (5.0) | 99 (4.2) | 359 (4.6) |
| Lobignonao | 91 (3.2) | 73 (2.8) | 88 (3.7) | 252 (3.2) |
| Moulé | 87 (3.0) | 89 (3.4) | 100 (4.3) | 276 (3.5) |
| Nouvielgane | 87 (3.0) | 88 (3.3) | 89 (3.8) | 264 (3.4) |
| Niombouna | 94 (3.3) | 107 (4.1) | 76 (3.2) | 277 (3.5) |
| Niombripo | 130 (4.6) | 122 (4.7) | 99 (4.2) | 351 (4.5) |
| Niaba | 138 (4.9) | 94 (3.6) | 87 (3.7) | 319 (4.1) |
| Nipodja | 76 (2.7) | 115 (4.4) | 85 (3.6) | 276 (3.5) |
| Ouidiaro | 116 (4.1) | 107 (4.1) | 78 (3.3) | 301 (3.9) |
| Palembra | 70 (2.5) | 72 (2.8) | 72 (3.1) | 214 (2.7) |
| Perbalembiro | 117 (4.1) | 87 (3.3) | 63 (2.7) | 267 (3.4) |
| Sarambour | 128 (4.5) | 101 (3.9) | 86 (3.7) | 315 (4.0) |
| Sidmoukar | 129 (4.5) | 90 (3.4) | 103 (4.4) | 322 (4.1) |
| Sinkiro | 69 (2.4) | 87 (3.3) | 54 (2.3) | 210 (2.7) |
| Soussoubro | 67 (2.3) | 62 (2.4) | 45 (1.9) | 174 (2.2) |
| Tordiero | 63 (2.2) | 63 (2.4) | 49 (2.1) | 175 (2.2) |
| Tiarkiro | 82 (2.9) | 77 (2.9) | 69 (2.9) | 228 (2.9) |
| Yelbelela | 85 (3.0) | 86 (3.3) | 65 (2.7) | 236 (3.0) |
| Yellela | 138 (4.8) | 121 (4.6) | 144 (6.1) | 403 (5.2) |
| **Sleep under bed net night**<br>**before survey** |  |  |  |  |
| No | 135 (4.8) | 462 (17.8) | 1574 (67.4) | 2171 (28.0) |
| Yes | 2704 (95.2) | 2132 (82.2) | 763 (32.6) | 5599 (72.0) |

**Table 2.** Demographical characteristics of the study participants to the passive survey.

|  | N | % |
| --- | --- | --- |
| **Sex** |  |  |
| Male | 2406 | 49.9 |
| Female | 2408 | 50.1 |
| **Age group** |  |  |
| < 6 months | 1240 | 25.7 |
| 6 - 59 months | 1400 | 29.1 |
| 5 - 9 years | 426 | 8.8 |
| 10 - 17 years | 272 | 5.6 |
| > 17 years | 1476 | 30.6 |
| **Primary Health Center (Target villages)** |  |  |
| Bondigui (*Kpedia*) | 134 | 2.8 |
| Diebougou (*Niaba*) | 57 | 11.8 |
| Dioura (*Tordiero, Sinkiro*) | 487 | 10.1 |
| Dolo (*Nouvielgane*) | 111 | 2.3 |
| Iolonioro (*Dontelo, Doumbourou, Kpakbalo, Niombouna, Ouidiaro, Perbalembiro, Sidmoukar, Tiakiro*) | 850 | 17.6 |
| Konsabla (*Nipodja*) | 211 | 4.4 |
| Loto (*Moulé*) | 83 | 1.7 |
| Niceo (*Bohéro*) | 218 | 4.5 |
| Pokouro (*Gongombiro, Sarambour, Niombripo*) | 1304 | 27.1 |
| Saptan (*Diagnon, Sousoubro*) | 185 | 3.8 |
| Tenguera (*Palembra*) | 123 | 2.5 |
| Tiankoura (*Dangbara, Kouloh, Lobignonao, Yelbelela*) | 664 | 13.8 |
| Tioyo (*Yellela*) | 387 | 8.1 |

### Prevalence of parasitemia and analysis of potential risk factors

A total of 2839 (70.5%), 2594 (64.4%) and 2337 (58%) participants were respectively examined in survey 1, survey 2 and survey 3. The proportion of participants identified at the time of the survey as symptomatic (fever or history of fever with RDT+) decreased from the first to the third survey; it was significantly higher during the first round of survey (39.9%) compared to the second (29.5%; *p* = 0.001) and third surveys (18.4%; *p* = 0.005). The prevalence of malaria infection on microscopy was high in all surveys (above 30%) with no difference between the first two surveys and a significantly lower for the third survey (*p* < 0.001): 41.5% (1178/2 839) in September 2016, 43.5% (1128/2 594) in December 2016 and 32.3% (756/2337) in June 2017. The mean parasite density in positive blood smear decreased progressively with 5801 asexual forms per μL of blood (95%CI 5034 - 6568) in the first survey, 1947 (95%CI 1675 - 2219) in the second and 1 780 (95%CI 1319 - 2241) in last survey (p < 0.001). Based on the RDT data, malaria prevalence was 38.2% (95%CI 36.4 - 40.2), 24.3% (95%CI 22.7 - 26.1) and 9.1% (95%CI 8.1 - 10.3) respectively in survey 1, survey 2 and survey 3 (*p* < 0.001).

In the univariate analysis of clinical malaria, the variables as the period of survey, gender, age, locality and sleeping under bed net the day before the survey were associated with the risk of malaria infection and then considered for the multivariate modeling (**Table 3**).

**Table 3.** Malaria prevalence and related risk factors during active surveys.

| **Risk factors** | **Cases (n/N)** | **Prevalence** | **Crude OR**<br>**(95%CI)** | ***p*** | **Adjusted OR**<br>**(95%CI)** | ***p*** |
| --- | --- | --- | --- | --- | --- | --- |
| **Survey** |  |  |  |  |  |  |
| Sept. 2016 (CSS1) | 1178/2839 | 41.5 | 1 |  | 1 |  |
| Dec. 2016 (CSS2) | 1128/2594 | 43.5 | 1.08 (0.97 - 1.20) | 0.1380 | 1.11 (0.99 - 1.24) | 0.053 |
| June 2017 (CSS3) | 756/2337 | 32.3 | 0.67 (0.60 - 0.75) | < 0.001 | 0.69 (0.60 - 0.80) | <0.001 |
| **Sexe** |  |  |  |  |  |  |
| Male | 1559/3765 | 41.4 | 1 |  | 1 |  |
| Female | 1503/4005 | 37.5 | 0.85 (0.77 - 0.93) | 0.0005 | 0.86 (0.79 - 0.95) | 0.004 |
| **Age** |  |  |  |  |  |  |
| 6 - 59 months | 1007/2370 | 42.5 | 1 |  | 1 |  |
| 5 - 9 years | 1323/3083 | 42.9 | 1.01 (0.91 - 1.13) | 0.7541 | 0.98 (0.88 - 1.10) | 0.789 |
| 10 - 17 years | 732/2317 | 31.6 | 0.62 (0.55 - 0.70) | < 0.001 | 0.58 (0.51 - 0.66) | <0.001 |
| **Village** |  |  |  |  |  |  |
| Bohero | 98/358 | 27.4 | 1 |  | 1 |  |
| Dangbara | 99/236 | 41.9 | 1.91 (1.34 - 2.72) | 0.0002 | 2.12 (1.48 - 3.01) | <0.001 |
| Diagno | 82/233 | 35.2 | 1.44 (1.00 - 2.05) | 0.0437 | 1.44 (1.01 - 2.07) | 0.043 |
| Dombourou | 158/345 | 45.8 | 2.24 (1.62 - 3.08) | <0.001 | 2.38 (1.73 - 3.28) | <0.001 |
| Dontélo | 119/289 | 41.2 | 1.85 (1.33 - 2.59) | 0.0002 | 2.07 (1.48 - 2.89) | <0.001 |
| Gongombiro | 171/442 | 38.7 | 1.67 (1.23 - 2.26) | 0.0008 | 1.68 (1.24 - 2.28) | 0.001 |
| Kouloh | 132/301 | 43.8 | 2.07 (1.48 - 2.88) | <0.001 | 2.08 (1.49 - 2.89) | <0.001 |
| Kpalbalo | 145/347 | 41.8 | 1.90 (1.38 - 2.62) | 0.0001 | 1.99 (1.44 - 2.75) | <0.001 |
| Kpédia | 103/359 | 28.7 | 1.06 (0.77 - 1.47) | 0.6950 | 1.04 (0.75 - 1.45) | 0.796 |
| Lobignonao | 127/252 | 50.4 | 2.69 (1.90 - 3.82) | <0.001 | 2.98 (2.11 - 4.21) | <0.001 |
| Moulé | 94/276 | 34.1 | 1.37 (0.97 - 1.92) | 0.0696 | 1.43 (1.01 - 2.02) | 0.042 |
| Nouvelgane | 79/264 | 29.9 | 1.13 (0.79 - 1.61) | 0.4864 | 1.23 (0.86 - 1.77) | 0.238 |
| Niombouna | 72/277 | 26.0 | 0.93 (0.65 - 1.32) | 0.6968 | 0.97 (0.67 - 1.39) | 0.882 |
| Niombripo | 158/351 | 45.0 | 2.17 (1.57 - 2.98) | <0.001 | 2.22 (1.61 - 3.05) | <0.001 |
| Niaba | 149/319 | 46.7 | 2.32 (1.67 - 3.22) | <0.001 | 2.21 (1.60 - 3.05) | <0.001 |
| Nipodja | 112/276 | 40.6 | 1.81 (1.29 - 2.53) | 0.0005 | 1.83 (1.30 - 2.57) | <0.001 |
| Ouidiaro | 120/301 | 39.9 | 1.75 (1.26 - 2.44) | 0.0007 | 1.77 (1.26 - 2.47) | 0.001 |
| Palembra | 103/214 | 48.1 | 2.46 (1.71 - 3.54) | <0.001 | 2.72 (1.89 - 3.90) | <0.001 |
| Perbalembiro | 142/267 | 53.2 | 3.01 (2.13 - 4.26) | <0.001 | 3.01 (2.15 - 4.23) | <0.001 |
| Sarambour | 89/315 | 28.2 | 1.04 (0.74 - 1.46) | 0.7995 | 1.02 (0.72 - 1.44) | <0.889 |
| Sidmoukar | 122/322 | 37.9 | 1.61 (1.16 - 2.24) | 0.0035 | 1.82 (1.31 - 2.54) | <0.001 |
| Sinkiro | 72/210 | 34.3 | 1.38 (0.95 - 2.00) | 0.0828 | 1.27 (0.87 - 1.84) | 0.200 |
| Soussoubro | 66/174 | 37.9 | 1.62 (1.10 - 2.38) | 0.0135 | 1.54 (1.04 - 2.28) | 0.028 |
| Tordiero | 87/175 | 49.7 | 2.62 (1.78 - 3.86) | <0.001 | 2.70 (1.84 - 3.96) | <0.001 |
| Tiarkiro | 63/228 | 27.6 | 1.01 (0.69 - 1.46) | 0.9458 | 1.07 (0.74 - 1.57) | 0.693 |
| Yelbelela | 108/236 | 45.8 | 2.23 (1.57 - 3.18) | <0.001 | 2.22 (1.56 - 3.15) | <0.001 |
| Yellela | 192/403 | 47.6 | 2.41 (1.76 - 3.29) | <0.001 | 2.59 (1.90 - 3.53) | <0.001 |
| **Slept under bed net the day before the survey** |  |  |  |  |  |  |
| No | 732/2171 | 33.7 | 1 |  | 1 |  |
| Yes | 2330/5599 | 41.6 | 1.40 (1.26 - 1.55) | <0.001 | 1.12 (0.98 - 1.28) | 0.082 |

*n* = number of clinical malaria cases; *N* = total number of participants surveyed;
OR = odds ratio; CI = confidence interval; *p* = *p*-value

In the multivariate analysis, girls seemed to have a lower risk of malaria infection (OR = 0.86; 95%CI = 0.79 - 0.95; *p* = 0.004). Similarly, the risk of malaria infection was significantly lower in the survey 3 (June 2017) at the beginning of the rainy season (OR = 0.69; 95%CI = 0.6 - 0.8; *p* < 0.001) compared to the survey 1 (September 2016) which was performed during the rainy season. Children aged 6 to 59 months had a higher risk of malaria infection compared to those aged 10 to 17 years (OR =0.58; 95%CI = 0.51 - 0.66). Finally, the risk of malaria infection varied significantly according to the village of residence. Participants living in Lobignonao (OR = 2.98; 95%CI = 2.11 - 4.21; *p* < 0.001) and Perbalembiro (OR = 3.01; 95%CI = 2.15 - 4.23; *p* < 0.001) were three-fold more frequently infected than those living in Bohero.

### Passive case detection

Over the study period, data for 4814 patients were extracted for consultation in the primary health centers. The history or episodes of fever was reported in 3 489 individuals (72.5%) of consultations. Malaria was diagnosed in 3238 individuals reaching 67.2% of the patients using RDT (**Table 4**). Malaria incidence showed a seasonal distribution, with an overall incidence rate estimated at 414.3 per 1000 person-years and large differences between months, varying from 18.8 to 69 per 1000 person-months (**Figure 2**). The month with lowest incidence was January 2017 (18.8/1000 person-months) with high peak in August 2017 (69/1000 person-months). Data comparison revealed that the incidence rate of 6 to 59 months’ age group was significantly higher than that in the other groups (*p* <0.001).

**Figure 2.**
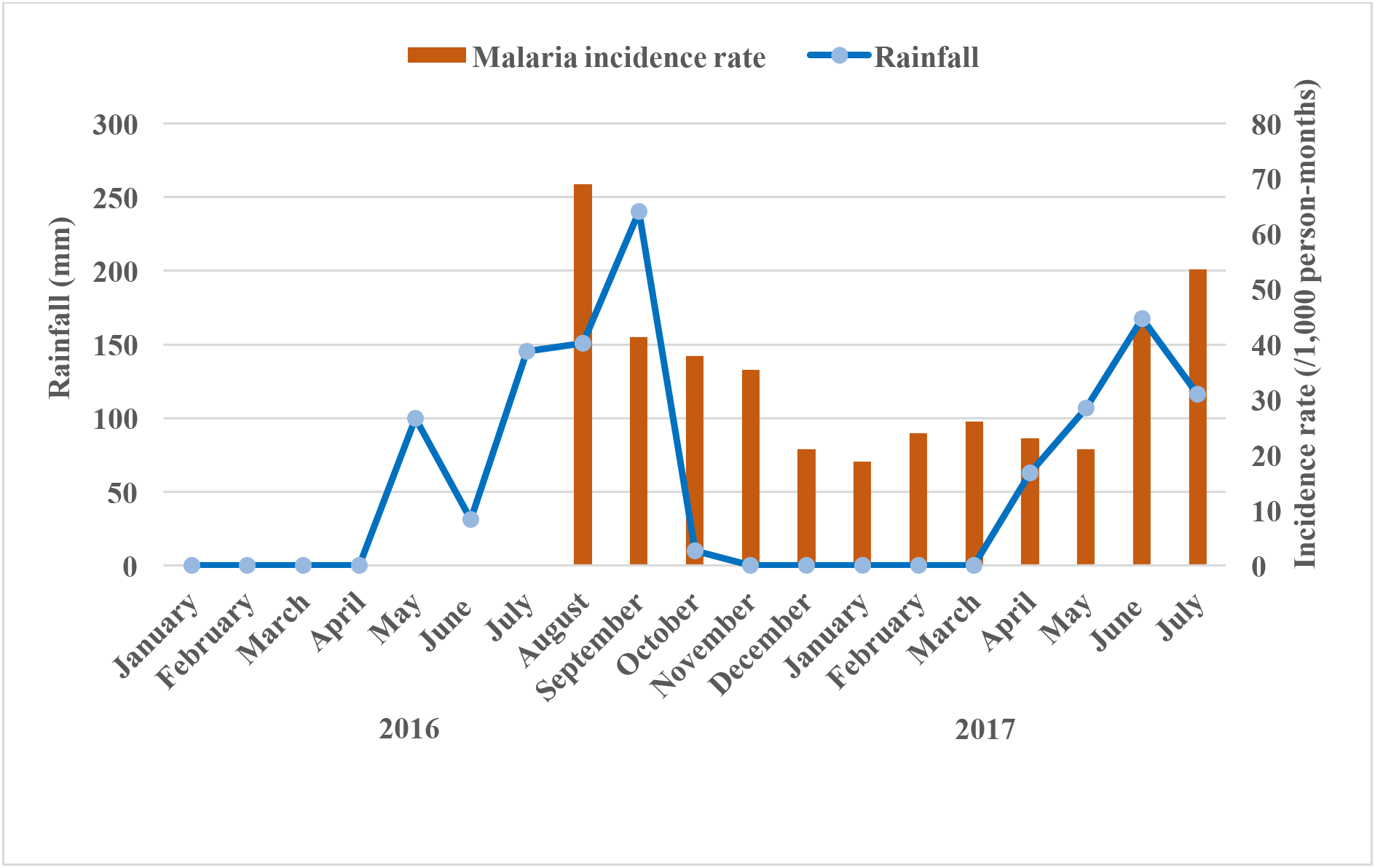
Monthly clinical malaria incidence during the study period as detected by passive case detection in primary health center and monthly cumulated rainfalls (data extracted from meteorological station of Diebougou).

**Table 4.** Data of individuals with suspected malaria fever and positive RDT during passive survey.

| Primary Health Centers | Consultations | Suspects cases<br>(History of fever) | Malaria Positive<br>(Positive RDT) |
| --- | --- | --- | --- |
| Bondigui | 134 | 108 (80.6%) | 106 (79.1%) |
| Diebougou | 57 | 38 (66.6%) | 31 (54.4%) |
| Diourao | 487 | 412 (84.6%) | 399 (81.9%) |
| Dolo | 111 | 82 (73.9%) | 71 (63.9%) |
| Iolonioro | 850 | 588 (69.1%) | 554 (65.2%) |
| Konsabla | 211 | 155 (73.4%) | 131 (62.1%) |
| Loto | 83 | 50 (60.2%) | 44 (53%) |
| Niceo | 218 | 184 (84.4%) | 183 (83.9%) |
| Pokouro | 1304 | 903 (69.2%) | 810 (62.1%) |
| Saptan | 185 | 141 (76.2%) | 131 (70.8%) |
| Tenguera | 123 | 97 (78.9%) | 91 (74%) |
| Tiankoura | 664 | 492 (74.1%) | 458 (69%) |
| Tioyo | 387 | 239 (61.7%) | 229 (59.2%) |
| **Total** | **4814** | **3489 (72.5%)** | **3238 (67.2%)** |

## Discussion

The strengths of the present study lay in the use of combination of a population-based data after an initial census and hospital data, the people investigated being residents of the study area and the assessment of malaria in different seasons. Moreover, the use of an active case detection strategy, which allows uncovering the real picture of malaria in the community [5]. In addition, the study used Giemsa microscopy as diagnostic tool for active case detection survey. This adds some reliability to our diagnosis as the technique is well appreciated for its properties in malaria case detection.

Our data showed a high burden of malaria, characterized by a prevalence of parasitemia in the active cases detection ranging from 32 - 43% during the transmission seasons with an overall incidence of malaria in the passive cases detection estimated to 414.3 per 1000 person-years. Data from previous reports and studies in the country [2, 6, 7] and other reports from sub-Saharan Africa [9–12] confirmed this high morbidity. Although incidence of malaria was passively detected in this study, there was a follow-up of infants through cross-sectional surveys and sensitization of the mothers for early health consultation. Early diagnosis and treatment of clinical malaria episodes had probably contributed to the reduction of severe malaria cases and related-mortality among the study population [13].

A seasonal distribution of malaria cases and infections was also observed, with the highest transmission period (coinciding with the wet season from June to November). In the cross-sectional surveys, cases were higher in September and December 2016. Clinical cases started raising in June with high peak in August. Such seasonal pattern of malaria was previously reported in Burkina Faso [14–16].

Children aged 6 to 59 months were the most affected by malaria both in active and passive cases detections confirming the high burden of malaria often notified within this age group [2, 17]. WHO recommends that children living in areas of the Sahel sub-region with higher seasonal malaria, benefit from seasonal malaria chemoprevention (SMC) [18]. In this regard, SMC (from August to November) in children aged 3-59 months can reduce consistently malaria incidence and burden during the rainy season [19].

Malaria prevalence varied significantly among villages. Participants living in Lobignonao and Perbalembiro were three-fold more infected than those living in Bohero. The variation of the malaria risk between neighboring villages sometimes separated by short distances, in endemic regions was documented by long times [20, 21]. In our study, this variation can be explained by several factors including access to health care, using rate of bed nets, local environment (proximity to mosquito breeding sites, vegetation, household structural features, …), variations in socio-demographic and socio-economic factors.

In Burkina Faso, malaria control policies include free distribution of LLINs, intermittent preventive treatment for pregnant women, treatment of malaria cases with artemisinin-based combination therapy, and SMC [22]. The malaria control measures evaluated in this study at individual level were the use of LLINs. The use of LLINs (sleeping under LLINs the night before the survey) was greater than 90% at the end of the rainy season but only 32% at the beginning of the rainy season. The LLINs using rate is similar to that observed in most of the West African countries [23–26]. Sleeping under LLINs seemed to be related to the nuisance caused by mosquitoes biting, when human populations were not bothered by mosquitoes, they did not use the treated nets [27–29].

We found no association between sleeping under LLINs the night before the survey and malaria prevalence. Although LLINs use plays a key etiologic role in malaria prevention in endemic countries, the use of LLINs were not significantly associated with participant’s malaria status. These results are in agreement with those of other authors, who also found no evidence of a lower odds of malaria infection among participants sleeping under LLINs in relative to those not [30–32]. This observed lack of association in our study could possibly be attributed to an inconsistent or inappropriate use of the nets or perhaps participants were exposed to mosquito bites during other times of the day or evening when the net was not in use. However, our findings conflict the results of other authors, who found an association between LLINs use and malaria infection [33–36].

## Conclusion

Malaria burden remains high in this region of Burkina Faso despite substantial efforts made in malaria control during the past decade. Diebougou health district is subjects to high burden of malaria with a seasonal variation in the prevalence of the disease. Differences was observed between communities among the prevalence of malaria and children under 5 years old were subject of malaria burden in this setting. This results reinforce the urgent need, while improving access to care and the using rate of LLINs, to develop alternative control strategies to complement those already existing.

## Data Availability

The datasets used during this article are available at the Institut de Recherche en Sciences de la Sante, Bobo-Dioulasso, Burkina Faso and will be made easily available on request, when required.

## Acknowledgements

This research was realized in the context of the REACT Project FSP/REFS N° 2006-22 supported by the INITIATIVE 5% and IRD. We thank the nurses and the biologist technicians who collected the data. We thank also the administration of the health district of Diebougou for their strong collaboration. We are grateful for all the inhabitants of the health district of Diebougou who took part in the surveys and participated actively in the data collection.

**Abbreviations**
ACT: Artemisinin-based Combination Therapy
CSS: Cross-Sectional Surveys
IRS: Indoor Residual Spray
ITNs: Insecticide-Treated Nets
LLINs: Long-Lasting Insecticidal nets
ODK: Open Data Kit
PHCs: Primary Health Centers
RCT: Randomized Controlled Trial
RDT: Rapid Diagnostic Test
SMC: Seasonal Malaria Chemoprevention

## Availability of data and materials

The datasets used and/or analysed during the current study are available at the Institut de Recherche en Sciences de la Santé, Bobo-Dioulasso, Burkina Faso and will be made easily available on request, when required.

## Authors’ contributions

IZ, CP, NM, RKD and AS conceived and designed the study. AS oversaw the surveys under the supervision of IZ, NM and RKD. AS and IZ analyzed the data. AS drafted the manuscript which was revised by the co-authors. All authors read and approved the final manuscript.

